# Serum myelin oligodendrocyte glycoprotein as an indicator of diagnosis and disease activity in multiple sclerosis

**DOI:** 10.1101/2025.09.06.25335236

**Authors:** Unsong Oh, Abena Kwegyir-Aggrey, Fatma Mufti, Caileigh Dintino, Stephanie Buxhoeveden, Julie McVoy, Ryan Canissario, Myla D. Goldman

**Affiliations:** Virginia Commonwealth University School of Medicine, Department of Neurology, Richmond, VA, U.S.A.; Accelerated Cure Project, Waltham, MA, U.S.A.

**Keywords:** MS, MOG, biomarker, blood, demyelination, disease activity

## Abstract

**Background:** Protein components of myelin, such as myelin oligodendrocyte glycoprotein (MOG), are potentially informative biomarkers of demyelination, but questions remain about the significance and usefulness of blood MOG protein levels in MS.

**Objectives:** In a case-control study, serum MOG protein levels were compared between MS and controls to determine the extent to which MOG levels are associated with the diagnosis and disease activity of MS.

**Results:** Serum MOG protein levels were higher in MS than healthy donors (mean difference 20.5 pg/ml, 95% CI 12.7, 28.2) and other neurological disease controls (mean difference 27.5 pg/ml, 95% CI 17.5, 40.5). Samples from MS in clinical relapse had higher MOG protein levels than MS in clinical remission (mean difference 15.3 pg/ml, 95% CI 6.11, 25.6). Correlation between serum MOG and NfL levels was moderate and MOG protein levels incrementally added information relevant to disease activity not captured by serum NfL levels.

**Conclusion:** MOG protein levels are elevated in the blood of individuals with MS and may be associated with disease activity.

## INTRODUCTION

MS is an inflammatory demyelinating disease of the central nervous system (CNS). Pathology is notable for varying degrees of demyelination and axonal injury. Whereas denuded axons from demyelination are considered the substrate for potential remyelination, axonal degeneration is considered irreparable. Biomarkers that are informative and discriminating with regards to the relative degrees of demyelination versus axonal injury associated with disease activity have the potential to improve prognostication, guide treatment decisions and enhance our understanding of disease mechanisms of MS.

MOG is a component of myelin enriched in the outer layer of myelin sheath [1]. MOG is considered to be primarily of CNS origin, and is therefore a potentially more specific marker of CNS demyelination than others, such as myelin basic protein (MBP) which are expressed in the CNS and the periphery [2]. While antibodies against MOG (i.e. anti-MOG IgG) have been well studied and shown to distinguish MOG antibody disease (MOGAD) from MS, less is known about the usefulness of MOG protein as a biomarker in MS.

We measured blood levels of MOG protein and assessed its potential as an indicator of diagnosis and disease activity in individuals with MS. To assess the significance of blood MOG protein levels, we tested the extent to which serum MOG levels (1) differentiate MS from controls, (2) associate with clinical relapse and (3) relate to serum neurofilament light chain (NfL) levels, a marker of neuroaxonal injury.

## METHODS

### Samples

#### Cohort 1. Accelerated Cure Project biorepository

(ACP biorepository). Serum samples from a cohort of participants with relapsing remitting MS (n = 33) and serum samples from age, sex and race-matched healthy donors (n = 35) were obtained from the Accelerated Cure Project biorepository. Subjects with MS were treatment-naive (i.e. not previously on disease modifying therapy at the time of sampling).

#### Cohort 2. Virginia Commonwealth University MS Biorepository

(VCU MS biorepository). Archived blood samples from subjects with treatment-naive relapsing MS (n = 30) and other neurological disease (OND) controls (n = 42) available from a local institutional review board-approved biorepository were used. OND controls included primary headache syndromes, dementia, motor neuron disease, neuropathy, and stroke. Subjects with MS were sampled at the time of diagnostic evaluation, met published diagnostic criteria [3] and had never been on disease modifying therapy. Subjects with primary headache syndromes had no neurological diagnoses other than headache. In a separate analysis, samples from an additional group of subjects with relapsing MS from the VCU MS biorepository were tested to compare MS in clinical relapse (n = 35) to those MS in clinical remission (n = 27). Clinical relapse was defined as the appearance of new or recurrent symptoms from MS lasting > 24 hours in individuals with at least 30 days of preceding neurologic stability, in the absence of fever and leading to a change in functional system scores on neurologist’s examination. Clinical remission was defined as at least 30 days of preceding neurological stability with stable functional system scores. All subjects provided written informed consent. Subjects were recruited for participation between June 2014 and 2024. Standard operating protocol was followed for collection and storage of blood samples. In short, serum was processed from peripheral blood obtained by venipuncture in serum separator tubes then aliquoted into cryovials and stored frozen at -80°C. Samples were thawed on ice at the time of the assays.

### Repeat freeze-thaw samples

Samples from 3 donors (cohort 2) were tested after repeat freeze-thaws to assess the stability of MOG protein levels in sera. After thawing on ice the sera were aliquotted, incubated at room temperature for one hour, and then returned to -80°C storage at least overnight. One aliquot from each set was subjected to the identical temperature cycle on day two and again on day three. On the day of assay the three 1X and 3X freeze-thawed aliquots were thawed on ice along with another previously unthawed aliquot of the same three sera and then assayed within one hour. Repeated measures one-way ANOVA test indicated no significant difference in MOG levels among samples from the same donor (F(1.4, 2.8) = 0.115, p = 0.832), indicating no significant impact of repeat freeze-thaws (up to 3) on serum MOG protein levels (Supplemental Figure 1).

### Biomarkers

MOG protein levels were measured using a commercially available human MOG ELISA kit (Abcam, #ab278115) according to the manufacturer’s protocol. Briefly, test samples or standards were incubated in solution with an anti-MOG antibody cocktail consisting of an affinity-tagged capture antibody and a horseradish peroxidase-conjugated detection antibody in microtiter wells precoated with an anti-affinity tag antibody to which the complexes became immobilized. After washing to remove unbound material, 3,3’,5,5’-tetramethylbenzidine development solution was added to the wells for colorimetric analysis performed on a plate reader (Synergy HT, Biotek). Each ELISA plate contained samples from MS and controls. Mean intra-assay coefficient of variation (CV) was 3.4% and mean inter-assay CV was 11.5% for the MOG ELISA. Serum Neurofilament light chain (NfL) was measured on the Simoa instrument platform (SR-X, Quanterix) using a commercially available kit (NF-light Advantage V2, Quanterix) according to the manufacturer’s protocol. Intra- and inter-assay CV was less than 10% for the NfL assay.

### Statistical analysis

Mean, standard deviation and 95% confidence intervals (95% CI) were determined for study parameters. Proportions (%) were determined for categorical variables. Estimation statistics [4] were used to determine the 95% confidence interval (CI) of the difference in means between groups. For p-value determinations, Mann-Whitney test of significance was used for two-group comparisons. Kruskal-Wallis with Dunn’s post-hoc comparisons was used for multiple group comparisons. Fisher’s exact test was used to compare categorical variables. Receiver operating characteristic (ROC) curve analysis was performed. Pearson correlation analysis was done for log transformed data. Multiple linear regression was used to adjust for covariates in the analysis of cohort 2 data comparing MS and OND. Z-scores adjusted for age and BMI were available for blood NfL [5] but not MOG levels. Therefore, standard z-scores were defined as z = (x-μ)/σ, where x is the measured biomarker level, μ and σ are the healthy donor mean and standard deviation respectively. Estimation statistics were performed on the Estimation Stats Web App (estimationstats.com)[4]. GraphPad Prism software was used for all statistical analyses other than estimation statistics.

## RESULTS

### Blood MOG protein levels are higher in MS compared to controls

We measured serum MOG levels from cohort 1 samples, comparing individuals with treatment-naive relapsing remitting MS to age, sex and race-matched healthy donors. Demographics of cohort 1 donors and biomarker levels for the groups are shown in Table 1. Serum MOG protein levels from individuals with MS were significantly higher than those of healthy donors (Figure 1A), with a mean difference of 20.5 pg/ml (95% CI 12.7, 28.2). Serum NfL levels were also significantly higher for MS compared to healthy donors (Table 1). ROC analysis was performed to assess to what extent biomarker levels discriminate between MS and healthy donors. ROC analysis of serum MOG protein levels for MS vs. healthy donors showed an area under the curve (AUC) of 0.85 (95% CI 0.75 to 0.95, p < 0.0001), indicating good discrimination between MS and controls (Figure 1B ). Similarly, ROC analysis showed good discrimination between MS and healthy donors based on serum NfL levels (AUC of 0.87, 95% CI 0.77 to 0.97, p < 0.0001).

**Figure 1.**
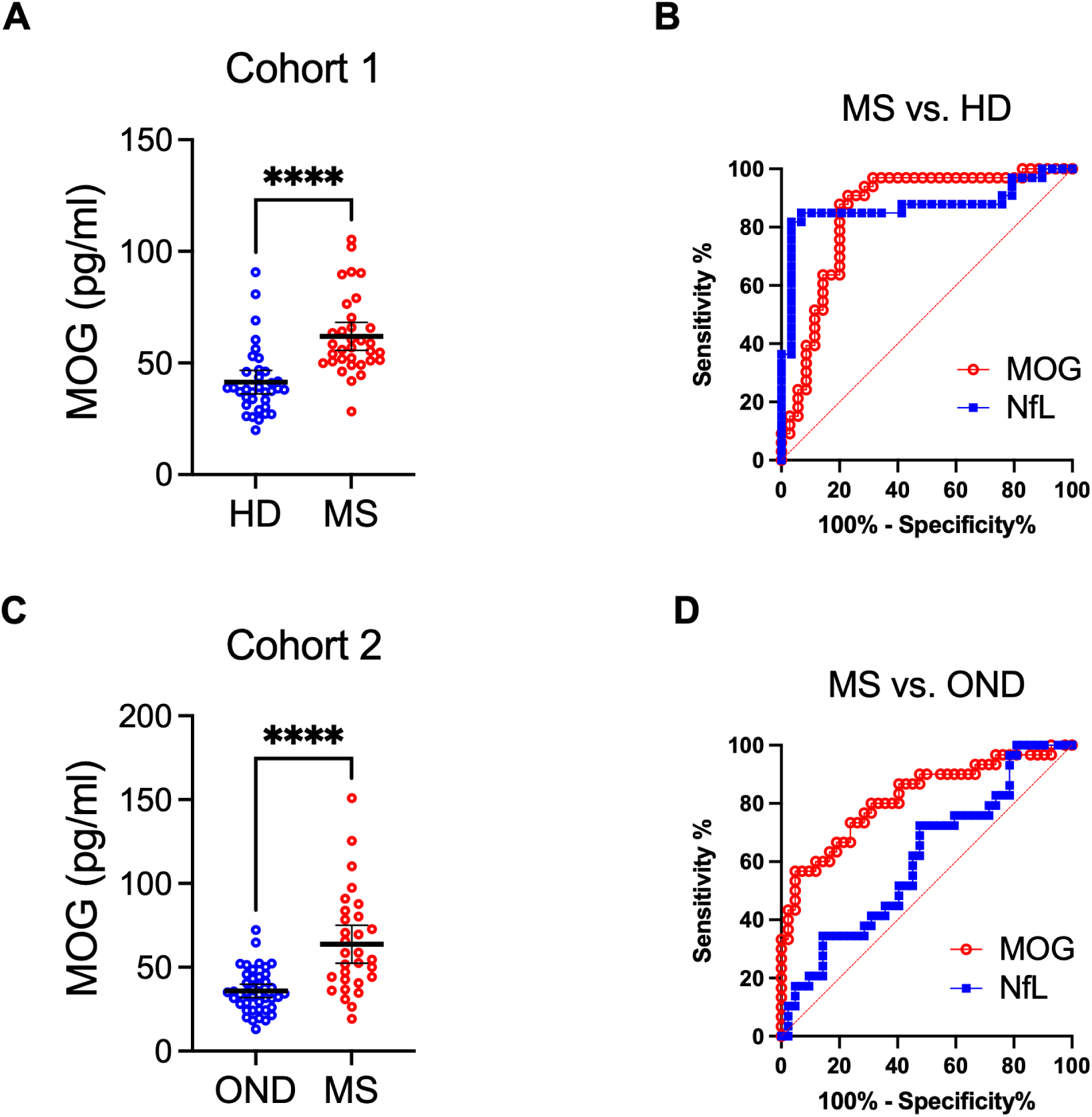
Blood MOG protein levels are higher in MS compared to controls. A) Cohort 1 (ACP biorepository) serum MOG levels for MS compared to healthy donors (HD). Mean and 95% confidence interval shown as line and error bars. **** p < 0.0001, Mann-Whitney test. B) ROC curve of serum MOG levels to classify MS vs. HD. C) Cohort 2 (VCU MS biorepository) serum MOG levels from individuals with MS compared to those from other neurological disorders (OND). Mean and 95% confidence interval shown as line and error bars. **** p < 0.0001, Mann-Whitney test. D) ROC curve of serum MOG levels to classify MS vs. OND.

**Table 1.** Demographics of cohort 1 (ACP biorepository) and biomarker levels.

|  | healthy donors<br>(n = 35) | MS<br>(n = 33) | p-value |
| --- | --- | --- | --- |
| Age, mean (SD) | 30.8 (4.7) | 32.9 (5.7) | 0.094 |
| Sex, female | 48.60% | 51.50% | >0.999 |
| Race (white) | 100% | 100% | >0.999 |
| BMI, mean (SD) | 28.3 (6.1) | 25.6 (5.5) | 0.1095 |
| Acute corticosteroid treatment* | NA | 8 (24%) | NA |
| On disease modifying therapy | NA | 0 | NA |
| serum MOG pg/ml, mean (SD) | 41.5 (15.3) | 62.0 (17.6) | <0.0001 |
| serum NfL pg/ml, mean (SD) | 6.9 (6.9) | 34.5 (79.5) | <0.0001 |

P-value: Mann-Whitney for continuous and Fisher's exact test for categorical variables. MS, multiple sclerosis. SD, standard deviation. BMI, body mass index. NA, not applicable. MOG, myelin oligodendrocyte glycoprotein. NfL, neurofilament light chain.
\*High dose corticosteroids received within 30 days prior to blood sampling.

Samples from a second, independent cohort of individuals with MS and OND controls were tested for serum MOG protein levels. Demographics of cohort 2 donors and biomarker levels for the groups are shown in Table 2 (and Supplemental Table 1 for demographics and biomarker levels by diagnosis). As shown in Figure 1C (and Supplemental Figure 2), serum MOG protein levels were significantly higher in MS compared to OND controls, with a mean difference of 27.5 pg/ml (95% CI 17.5, 40.5). NfL levels, however, did not differ significantly between MS and OND controls (Table 2). ROC analysis of serum MOG protein levels for MS and OND controls showed an AUC of 0.82 (95% CI 0.72 to 0.92, p < 0.0001), indicating good discrimination between MS and OND controls (Figure 1D). By comparison, ROC analysis showed poor discrimination between MS and OND based on NfL levels (AUC 0.61, 95% CI 0.48 to 0.74, p = 0.125). Cohort 2 cases and controls were not as well matched (Supplemental Table 1), owing to inherent diversity in the demographics of the different diseases. Therefore, multiple linear regression was performed to test whether the diagnosis of MS significantly predicted MOG levels, adjusting for covariates. Diagnosis of MS remained significantly associated with higher MOG levels (estimate 24.8; 95% CI 14.2, 35.4; p < 0.0001) after adjusting for age, sex, race and BMI (Supplemental Table 2).

**Table 2.** Demographics of cohort 2 (VCU MS biorepository) and biomarker levels.

|  | OND<br>(N = 42) | MS<br>(N = 30) | p-value |
| --- | --- | --- | --- |
| Age, mean (SD) | 46.3 (14.4) | 40.0 (12.3) | 0.085 |
| Sex, female (%) | 65.9 | 63.3 | >0.999 |
| Race, % White:Black:Other | 48:52:0 | 50:43:6 | 0.285 |
| BMI, mean (SD) | 31.9 (7.1) | 30.0 (7.3) | 0.281 |
| Acute corticosteroid treatment* | NA | 6 (22%) | NA |
| On disease modifying therapy | NA | 0% | NA |
| Serum MOG pg/ml, mean (SD) | 35.9 (12.8) | 63.8 (30.2) | <0.0001 |
| Serum NfL pg/ml, mean (SD) | 36.0 (159.8) | 18.3 (17.1) | 0.127 |

P-value: Mann-Whitney for continuous and Fisher's exact test for categorical variables. OND, other neurological disorders. MS, multiple sclerosis. SD, standard deviation. BMI, body mass index. NA, not applicable. MOG, myelin oligodendrocyte glycoprotein. NfL, neurofilament light chain. \*High dose corticosteroids received within 30 days prior to blood sampling.

MOG protein levels were also measured in the cerebrospinal fluid (CSF) from cohort 2 donors that had paired CSF and serum samples available. There was a moderate correlation between cerebrospinal fluid and serum MOG protein levels (Pearson r = 0.484, p = 0.005, Supplemental Figure 3).

### Blood MOG protein levels are associated with clinical disease activity in MS

In a separate analysis, samples from an additional group of subjects with relapsing MS available from the VCU MS biorepository were tested to assess the utility of blood MOG protein levels as a biomarker of disease activity in MS. We compared MOG protein levels in samples from individuals with MS experiencing a clinical relapse at the time of blood sampling (MS in relapse) to those in clinical remission (MS in remission). Demographics of the cohort and biomarker levels for cases and controls are shown in Table 3. As shown in Figure 2A, serum MOG protein levels for MS in relapse were significantly higher than those of MS in remission, with a mean difference of 15.3 pg/ml (95% CI 6.11, 25.6). Serum NfL levels were also higher for MS in relapse compared to MS in remission (Table 3). When using a cutoff of 2 standard deviations above the healthy donor mean for MOG (i.e. 72.1 pg/ml), 28.6% of MS in relapse samples were above the MOG cutoff. By comparison, when using the cutoff of 2 standard deviations above healthy donor mean for NfL (i.e. 9.8 pg/ml), 48.6% of the MS in relapse samples were above the cutoff. When using either the MOG or the NfL cutoff, 57% of MS in relapse samples were above the cutoff, indicating that the use of both MOG and NfL levels identify a higher proportion of MS in relapse than either alone. MOG but not NfL was high in 8.6% of MS in relapse (Figure 2B, quadrant a), and NfL but not MOG levels was high in 28.6% of MS in relapse (Figure 2B, quadrant c). These results suggest that serum MOG levels provide incremental additional knowledge not captured by NfL regarding disease activity in MS.

**Figure 2.**
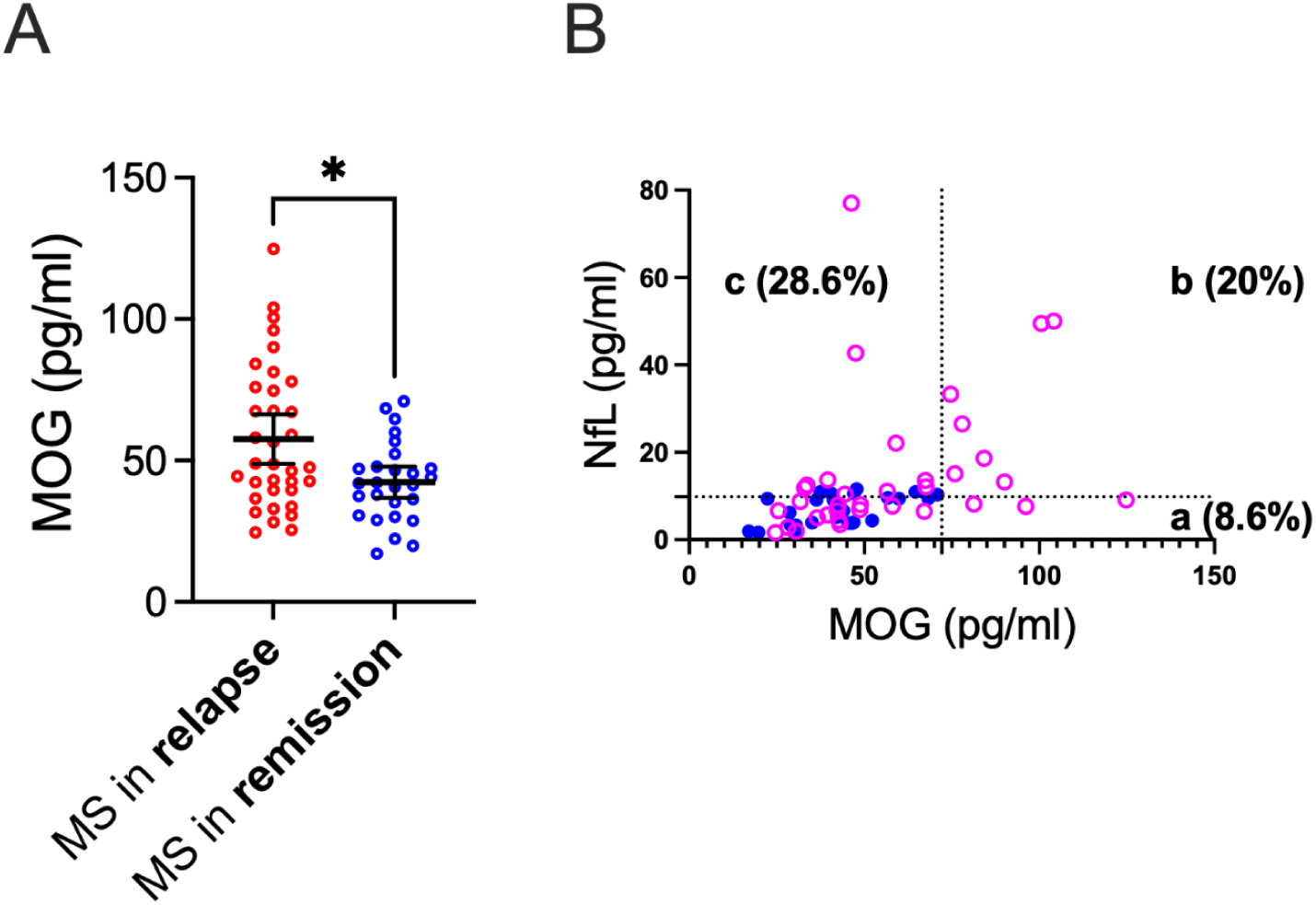
Blood MOG protein levels are associated with clinical disease activity in MS. A) Serum MOG protein levels for individuals with MS in clinical relapse (MS in relapse) compared to those in remission (MS in remission). Mean and 95% confidence interval for serum MOG shown as line and error bars. * p < 0.05, Mann-Whitney test. B) NfL vs. MOG levels for MS in relapse (open circles) and MS in remission (filled circles). Dotted lines indicate cutoff values of 2 standard deviations above healthy donor means for MOG (X-axis) and NfL (Y-axis).

**Table 3.** Demographics of MS in clinical relapse and remission (VCU MS biorepository) and biomarker levels.

|  | MS in relapse<br>(N = 35) | MS in remission<br>(N = 27) | p-value |
| --- | --- | --- | --- |
| Age, mean (SD) | 37.9 (9.7) | 39.2 (9.2) | 0.444 |
| Sex, female | 80.0% | 81.5% | >0.999 |
| Race, % White:Black | 80:20 | 70.4:29.6 | 0.551 |
| BMI, mean (SD) | 31.2 (8.1) | 31.4 (6.8) | 0.67 |
| Acute corticosteroid treatment* | 0 | 0 | NA |
| On disease modifying therapy | 60% | 80% | 0.053 |
| serum MOG pg/ml, mean (SD) | 57.6 (25.3) | 42.3 (14.0) | 0.019 |
| Serum NfL pg/ml, mean (SD) | 15.5 (16.4) | 7.0 (3.4) | 0.017 |

P-value: Mann-Whitney for continuous and Fisher's exact test for categorical variables. MS, multiple sclerosis. BMI, body mass index. SD, standard deviation. NA, not applicable. MOG, myelin oligodendrocyte glycoprotein. NfL, neurofilament light chain. \*High dose corticosteroids received within 30 days prior to blood sampling.

### Correlation between MOG and NfL in MS

The relationship between MOG and NfL levels was assessed as exploratory surrogate markers for demyelination and axonal loss. Among all subjects with MS (cohort 1 and cohort 2), there was a moderate correlation between serum MOG and NfL levels (Pearson r = 0.523, p < 0.0001, Figure 3A). To explore the premise that blood MOG and NfL reflect demyelination and axonal loss, respectively, we examined MOG to NfL ratio for each subject as a measure of relative demyelination vs. axonal loss in a given individual with MS. We used standard z-scores to calculate MOG vs. NfL ratios given the difference in range for serum MOG (19.3 - 150.9 pg/ml ) and serum NfL (3.8 - 389.9 pg/ml) in subjects with MS. A frequency histogram was used to visualize the distribution of MOG/NfL z-score ratios among “active” MS samples defined as > 2 standard deviations above healthy donor means for MOG or NfL. As shown in Figure 3B, the distribution of MOG/NfL z-score ratios showed a single peak with a few outliers showing high MOG/NfL or low MOG/NfL ratios, suggesting that the majority of MS samples fall within a single homogenous population with respect to their MOG/NfL ratio.

**Figure 3.**
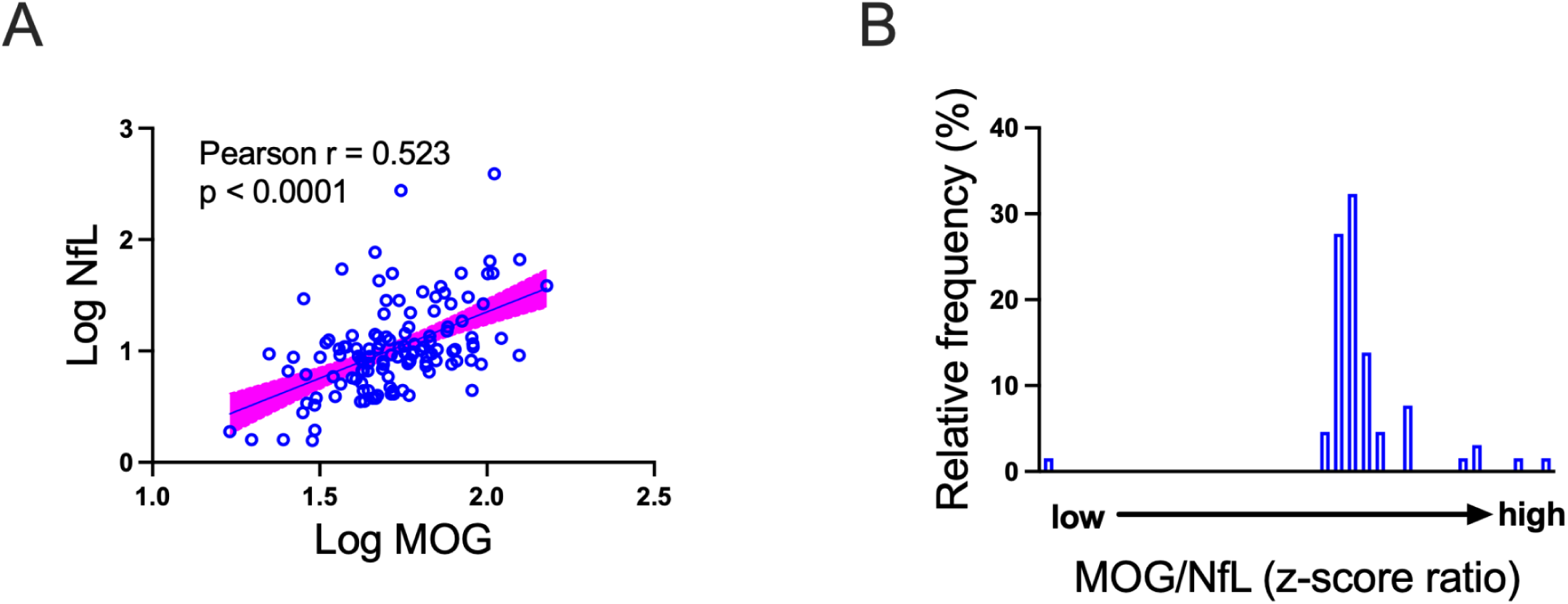
Correlation between MOG and NfL in MS. A) Log transformed serum MOG vs. log transformed serum NfL for all subjects with MS (cohort 1 and cohort. B) Frequency distribution histogram of MOG/NfL z-score ratios for subjects with MS with high MOG or high NfL levels (> 2 standard deviations above healthy donor mean). Bin size = 0.25.

## DISCUSSION

In a case-control study, we show that serum MOG protein levels are higher in those with MS compared to healthy donors and OND controls. ROC curve analysis indicates that serum MOG protein levels provide good discrimination between MS and controls. Serum MOG protein levels were also higher in samples from the cohort of MS patients in clinical relapse compared to those in remission, suggesting that elevated serum MOG levels are associated with disease activity in MS. These results indicate that serum MOG may be a useful biomarker for MS, likely representing the underlying pathology of demyelination.

Our results support the use of serum MOG protein levels in MS to complement existing tools such as blood NfL [6],[7]. We show that MOG protein levels are informative in several ways. When compared to NfL alone, the addition of MOG protein levels incrementally improved the identification of samples from MS in clinical relapse. Whereas 48.6% of MS in clinical relapse samples were identified by high NfL levels alone, 57% of MS in relapse were identified by the criteria of either high MOG or high NfL levels. This incremental improvement stemmed from a small proportion (8.6%) of MS in relapse that were identified by high MOG but not high NfL levels, suggesting that MOG captures additional information regarding MS disease activity not captured by NfL alone. The concept that NfL alone is inadequate to identify all disease activity in MS is supported by prior research showing that a substantial proportion of those with newly enhancing MS lesions on MRI did not have elevation in blood NfL levels [8]. Additionally, our results show that MOG levels were more discriminating compared to NfL in the context of OND, consistent with the knowledge that NfL levels are high in many neurological disorders [9]. Therefore, serum MOG levels may be particularly useful when comorbid neurological conditions are present and confound the interpretation of NfL in subjects with MS.

Based on the premise that MOG protein and NfL are respectively the byproducts of demyelination and axonal injury, one could ask what MOG and NfL levels tell us about the relative degrees of demyelination and axonal injury resulting from MS disease activity. We found a moderate correlation between serum MOG and NfL levels, suggesting that demyelination and axonal injury are often linked by the same underlying pathobiology. Of interest to us were the outliers in the MOG versus NfL correlation analysis which showed either high MOG/low NfL or low MOG/high NfL (i.e. quadrants **a** and **c** of Figure 2B). One hypothesis might be that these outliers represent MS endophenotypes defined by those with a predominantly demyelinating (high MOG/low NfL) lesion phenotype versus those with a predominantly neurodegenerative (low MOG/high NfL) lesion phenotype. Alternatively, it may be that MOG/NfL ratio reflects the temporal dynamics of lesion evolution in MS where the initial demyelinating injury sheds MOG protein from the outer, “exposed”, part of myelin, with subsequent axonal transection/degeneration releasing NfL. The frequency distribution of MOG/NfL ratios (Figure 3B) appears to show that the majority of the samples would likely exhibit a mixed pathology of demyelination and axonal injury. Whether the few outliers represent the extreme points along lesion evolution or represent durable lesion endophenotypes (predominantly demyelinating or predominantly neurodegenerative) cannot be answered by our data. Interestingly, a recent report showed that the peak of blood NfL levels lag the onset of MS lesion formation by a median of 8 weeks [8]. Also of interest is the report that showed that a significant increase in blood MOG levels precede the increase in blood NfL in samples from pre-symptomatic MS [10]. Our guess is that peak of blood MOG levels occurs early in MS lesion formation, followed by the later peak of serum NfL levels, and that differences in relative MOG vs. NfL levels largely reflect timing of blood sampling relative to lesion formation. Future prospective studies are needed to clarify to what extent biomarker levels reflect the temporal dynamics and the pathology of lesion formation in MS.

The main limitation of this study is the retrospective case-control design. The archived blood samples were obtained from two biorepositories. Although possible, differences in sample processing are unlikely to explain these results given similar standard operating procedures and the inclusion of internal controls in each cohort. We did not match age, sex or BMI for OND controls compared to MS (Supplemental Table 1) due to the inherent diversity in the demographics of the different diseases, with the OND group being older, in particular. Nevertheless, multiple linear regression modeling showed that the diagnosis of MS was significantly associated with higher blood MOG protein levels compared to OND after adjusting for covariates such as age, sex, race and BMI. Age- and BMI-adjusted z-scores were available for NfL [5] but not MOG. We did not have the appropriate sample size of healthy donors to adequately assess the effect of relevant biological variables such as age, sex and BMI on blood MOG protein levels. Therefore, we used the standard z-scores (i.e. distance from healthy donor mean expressed in units of standard deviations) for MOG and NfL for within-person MOG/NfL ratio calculations. We did not test samples from other CNS inflammatory demyelinating disorders such as neuromyelitis optica spectrum disorder or MOG antibody disease. Future prospective studies are needed to further establish MOG protein as a useful biomarker for diagnosis, prognosis and monitoring response to therapy in MS and other CNS inflammatory demyelinating disorders.

In summary, our results support the use of blood MOG protein as an informative biomarker of MS diagnosis and disease activity. These results add to the knowledge guiding the clinician in the optimal use and interpretation of biomarker panels for MS that include MOG.

## Data Availability

All data produced in the present study are available upon reasonable request to the authors

## Statements and declarations

This study was approved by the Virginia Commonwealth University Institutional Review Board (protocol #HM14322). Participants provided written informed consent. The authors declared no potential conflicts of interest with respect to the research, authorship, and/or publication of this article. This work was supported by philanthropic funding from the ZiMS Fund and the Mitchell Family Fund.

---

**Supplemental Table 1.** Demographics of cohort 2 (VCU MS biorepository) and biomarker levels by diagnosis.

|  | headache<br>(n = 25) | dementia<br>(n = 4) | motor neuron<br>disease (n = 3) | neuropathy<br>(n = 6) | stroke (n = 4) | MS (n = 30) |
| --- | --- | --- | --- | --- | --- | --- |
| Age, mean (SD) | 37.9 (10.2) | 61.3 (10.6) | 57.3 (10.0) | 56.8 (10.8) | 54.5 (7.1) | 40.0 (12.3) |
| Sex, female | 88% | 75% | 33.3% | 0% | 25% | 63.3% |
| Race, % White:Black | 36:64 | 75:25 | 33.3:66.6 | 80:20 | 50:50 | 50:43 |
| BMI, mean (SD) | 34.5 (7.0) | 25.7 (7.1) | 27.3 (5.3) | 26.2 (1.7) | 30.9 (5.1) | 30.0 (7.3) |
| serum MOG pg/ml, mean<br>(SD) | 36.8 (13.4)* | 42.0 (8.1) | 22.1 (10.6)* | 29.8 (11.1)* | 42.4 (8.3) | 63.8 (30.2) |
| Serum NfL pg/ml, mean<br>(SD) | 6.2 (3.5)* | 21.7 (5.4) | 28.7 (15.6) | 187.2 (420.3) | 15.7 (12.4) | 18.3 (17.1) |
| serum NfL, z-score<br>(adjusted) | 0.3 (1.4)* | 1.9 (0.7) | 2.4 (0.3) | 1.5 (1.5) | 1.3 (1.3) | 1.5 (1.6) |

MS, multiple sclerosis. SD, standard deviation. BMI, body mass index. MOG, myelin oligodendrocyte glycoprotein. NfL, neurofilament light chain. Adjusted NfL z-scores were adjusted for sex and BMI [5].\* P < 0.05, Kruskal-Wallis with Dunn's post-hoc multiple comparisons vs. MS for MOG and NfL

**Supplemental Table 2.** Multiple linear regression model of MOG protein level for cohort 2 (MS and OND)

| Predictors | estimates | 95% CI | t | p-value |
| --- | --- | --- | --- | --- |
| (intercept) | 67.334 | 35.5 to 99.2 | 4.224 | < 0.0001 |
| diagnosis [multiple sclerosis] | 24.79 | 14.2 to 35.4 | 4.661 | < 0.0001 |
| age | -0.096 | -0.501 to 0.309 | 0.4753 | 0.6362 |
| race [Black] | -7.068 | -17.68 to 3.532 | 1.332 | 0.1876 |
| sex [male] | -12.68 | -24.40 to -0.962 | 2.161 | 0.0344 |
| body mass index | -0.5887 | -1.342 to 0.1646 | 1.561 | 0.1234 |
| model R<sup>2</sup> = 0.3883, F (6, 65) = 6.878, p < 0.0001 |  |  |  |  |

**Supplemental Figure 1.**
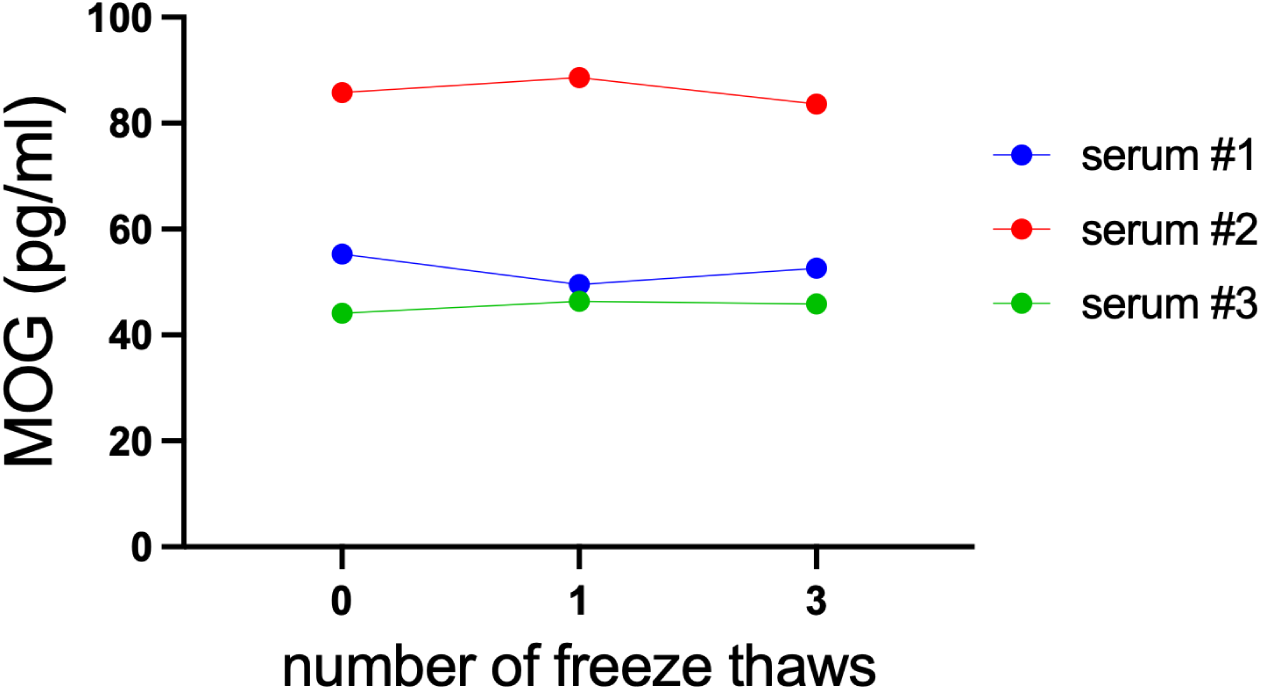
The effect of repeat freeze thaws on serum MOG protein level.

**Supplemental Figure 2.**
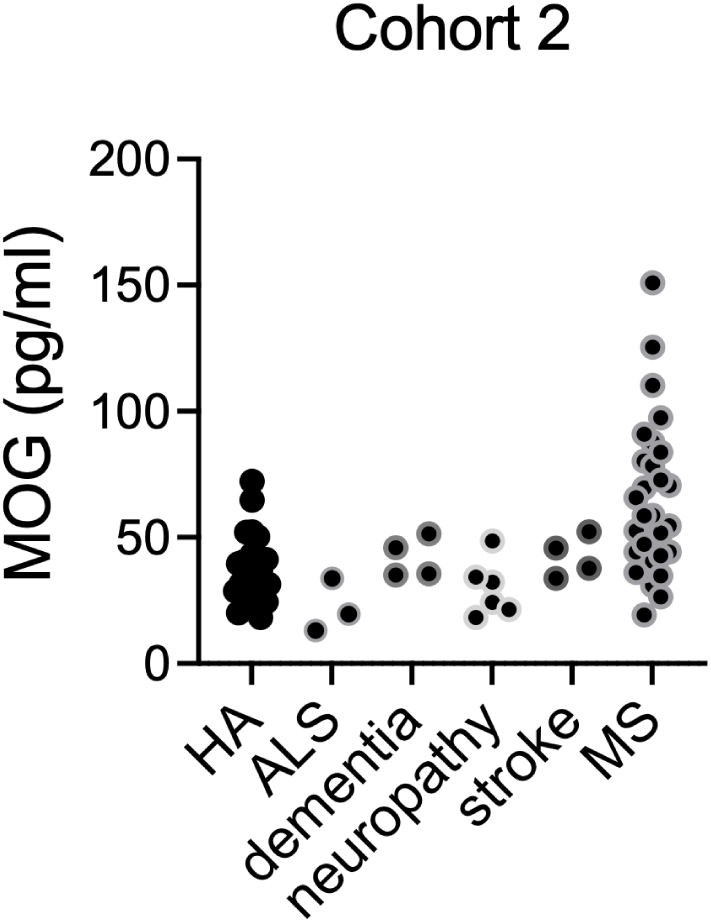
Blood MOG protein levels are higher in MS compared to controls. Cohort 2 (VCU MS biorepository) serum MOG levels for individuals with MS compared to those from individuals with primary headache syndromes (HA), motor neuron disease (ALS), dementia, neuropathy and stroke.

**Supplemental Figure 3.**
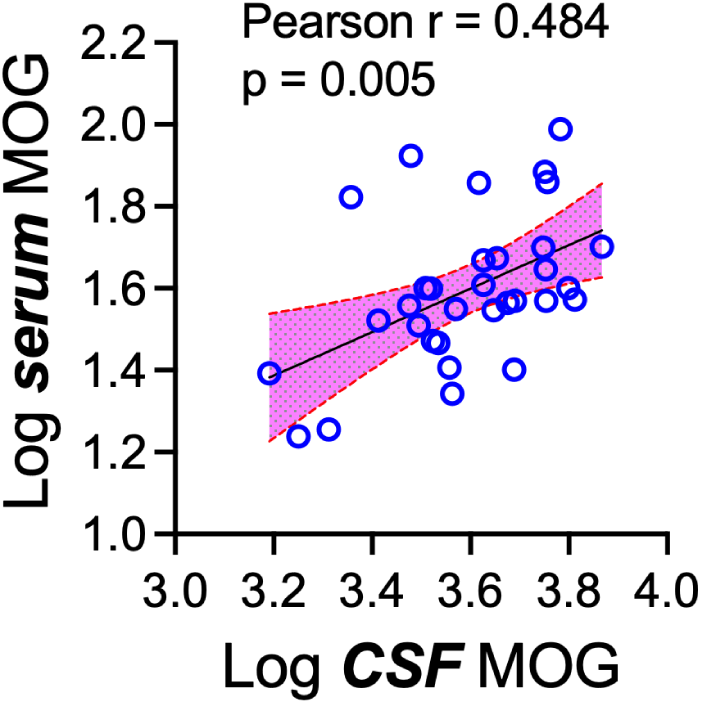
Correlation between serum and CSF MOG protein levels. Serum and CSF MOG protein levels were compared for cohort 2 samples for whom both serum and CSF samples were available. Shown are log transformed MOG levels.

